# Genetic and environmental architecture of violent victimisation across development and sex: A study of 4.5 million Nordic twins and siblings

**DOI:** 10.64898/2025.12.18.25342608

**Authors:** Amir Sariaslan, Ralf Kuja-Halkola, Jonas Forsman, Joonas Pitkänen, Ebba du Rietz, Zheng Chang, Brian M. D’Onofrio, Mikko Aaltonen, Henrik Larsson, Pekka Martikainen, Paul Lichtenstein, Seena Fazel

**Author notes:** All correspondence should be sent to: Dr Amir Sariaslan, Department of Psychiatry, University of Oxford, Warneford Hospital, Oxford OX3 7JX, United Kingdom. Competing interests: HL reports receiving grants from Takeda and Shire Pharmaceuticals; personal fees from and serving as a speaker for Medice, Shire/Takeda Pharmaceuticals and Evolan Pharma AB; all outside the submitted work. HL is editor-in-chief of JCPP Advances. ZC received lecture honoraria from Takeda Pharmaceuticals, outside the submitted work. Author contributions: AS and SF conceived and designed the study. AS conducted the statistical analyses, with assistance from RKH, and drafted the manuscript together with SF. All authors (AS, RKH, JF, JP, EdR, ZC, BMD, MA, HL, PM, PL, and SF) interpreted the findings, critically revised the draft, approved the final version of the manuscript, and take full responsibility for the accuracy and integrity of the work.

## Abstract

Violent victimisation affects 1-4% of populations annually and constitutes a major risk factor for psychiatric morbidity and suicidal behaviours. However, the aetiological mechanisms underlying vulnerability to severe victimisation remain poorly understood. We examined genetic and environmental contributions to victimisation risk across development using nationwide family data from 4,458,368 individuals born in Sweden (1973-2004) and Finland (1970-2003). Violent victimisation was identified through hospital admissions and mortality records. Quantitative genetic models estimated additive genetic, shared environmental, and unique environmental influences across developmental periods, with sex-limitation analyses examining sex-specific effects. Among 154,209 individuals (2.9%) with documented victimisation, familial aggregation was proportional to genetic relatedness (adjusted hazard ratios: 6.0 [95% CI 4.0-9.0] for monozygotic twins; 1.4 [95% CI 1.4-1.5] for paternal half-siblings). Aetiological architecture varied substantially across development. Childhood-onset victimisation showed high heritability (h^2^=70%, 95% CI 44-95%) with notable shared environmental contributions (c^2^=22%, 95% CI 9-35%). Adolescent-onset and adult-onset victimisation demonstrated lower heritability (h^2^=40-44%) with predominant unique environmental effects (e^2^=56-60%) and negligible shared environmental influence. Sex-limitation models revealed comparable heritability between sexes but moderate cross-sex genetic correlations (r_g_=0.77-0.78), indicating partially distinct genetic pathways. Violent victimisation therefore exhibits a developmentally dynamic genetic architecture, with heritability decreasing and unique environmental contributions increasing from childhood to adulthood. Partially sex-specific genetic pathways underscore the need for age- and sex-stratified genomic investigations.

---

Although violent victimisation prevention represents a major public health priority, there has been limited progress in developing effective preventive interventions. Representative victim surveys suggest that approximately 1-4% of populations experience violence annually, with rates varying by definition and methodology.^1–3^ Surveys in the US (2018-2022) have further found that around 0.3% experience aggravated assaults.^3^ Violent victimisation disproportionately affects marginalised communities, including low-income families,^4^ children placed in out-of-home care,^5^ and those with physical and cognitive disabilities.^6–10^ Exposure to violent victimisation is consistently associated with more than a doubling of the subsequent risks of long-term adverse outcomes, including psychiatric disorders, antisocial behaviours, and premature mortality.^11,12^ Understanding the aetiology of violent victimisation is therefore key to developing effective preventive interventions.

Behaviour genetic investigations have applied various twin,^13–16^ pedigree,^16,17^ adoption^18^ and genomic^19^ designs to understand the relative contributions of genetic and environmental determinants of violent victimisation. The role of genetic factors may appear counterintuitive, but such influences are largely expressed indirectly through behavioural and social pathways. For instance, perpetrators of violence tend to target those with neurodevelopmental disorders,^9^ including those with reduced cognitive functioning,^20^ and psychiatric disorders,^7,10^, traits that are at least moderately heritable. Similarly, individuals who engage in antisocial behaviours and substance misuse, also partially heritable traits, are more likely to seek out similar peers and self-select into criminogenic environments, which places them at a higher risk of being victimised. Reported heritability estimates of violent victimisation have typically ranged from 40% to 60% but have lacked precision due to small sample sizes and overreliance on parent- and self-reported measures. The same studies have typically found that relevant environmental influences tend to be individual-specific rather than shared within families, particularly after childhood.^21^ The lack of high quality data means that we do not know the degree to which aetiological contributions vary across developmental stages or simply reflect measurement differences.^21,22^ Moreover, sex differences in the genetic architecture of violent victimisation remain poorly understood, largely resulting from a lack of statistical power to compare same-sex and opposite-sex relatives.^16,23^

To address these gaps in the literature, we have examined the entire populations of Sweden and Finland born across three decades and identified twins and non-twin siblings with and without experiences of violent victimisation. To increase statistical power, we ran parallel analyses within each country and pooled the results. We primarily focused on victimisation events that required medical care and/or resulted in death, but we also examined police-reported violent victimisation events in Finland. We fitted quantitative genetic sex-limitation models, which allowed us to decompose the variance in violent victimisation risk across genetic and environmental components, and test for age and sex moderation effects.

## Methods

We used nationwide registers in Sweden and Finland containing individual-level health and sociodemographic data, linked within each country using personal identification numbers for accurate cross-register linkage. Access to pseudonymised administrative population data was approved by the Swedish Ethical Review Authority (Dnr 2020-06540; Dnr 2022-06204-02), the Ethics Board of Statistics Finland (TK-53-1490-18), and the Finnish Institute of Health and Welfare (THL/2180/14.02.00/2020). Informed consent is not legally mandated for research using national registers with pseudonymised identifiers in either country.

The National Patient Register, overseen by the Swedish National Board for Health and Welfare, and the Care Register for Health Care, managed by the Finnish Institute for Health and Welfare (THL), include data on inpatient hospitalisation episodes (Sweden: 1973-2020; Finland: 1969-2020) and specialist outpatient care (Sweden: 2001-2020; Finland: 1998-2020). Diagnoses were coded using the Swedish and Finnish adaptations of the International Classification of Diseases (ICD), covering the 8th to 10th revisions. Information on mortality, including primary and contributing causes of death, was sourced from each country’s Causes of Death Registers, which employed the same ICD coding as the hospital records. Sociodemographic data, including emigration dates, were obtained from a range of population administrative registers maintained by Statistics Sweden and Statistics Finland. Zygosity data used to identify monozygotic and dizygotic twins were derived from the Swedish Twin Register.

The study sample comprised all children born in Sweden (1973-2004; n=3,269,545) and Finland (1970-2003; n=2,103,835). We excluded individuals who could not be linked to their biological parents (n_Sweden_=42,340; n_Finland_=34,043), as this information was required to classify sibling relationships. Following these exclusions, the final analytical sample included 5,296,997 individuals.

### Assessment of violent victimisation

Consistent with earlier work,^7^ we defined violent victimisation as either an inpatient care episode, secondary care outpatient visit, or death associated with a diagnosis of an injury purposefully inflicted by another person (ICD-8/9: E960-E969; ICD-10: X85-X99 and Y00-Y09). To capture incidents that may not have required medical care, we also considered police-reported violent victimisation as a secondary outcome.

This included individuals recorded as victims of violent offences (e.g., attempted homicide, assault, robbery, unlawful threats, or sexual offences) in Finnish police reports from 2003 to 2020. The latter analyses were limited to individuals who were alive and residing in Finland as of the end of 2002 (n=2,015,805).

### Familial clustering

To investigate the extent to which violent victimisation clusters within families, we estimated associations between the risk of violent victimisation in probands and their co-relatives using Cox regression models, with attained age specified as the underlying time scale. The magnitude of the associations was expressed as hazard ratios and their uncertainty quantified using 95% confidence intervals. A detailed account of the approach is available elsewhere.^24^ In brief, we identified pairs of monozygotic and dizygotic twins in the Swedish sample, and all non-twin full siblings, maternal half-siblings, and paternal half-siblings in both countries. To maximise statistical power, all combinations of twin and sibling pairs were analysed twice, allowing each individual to contribute both exposure and outcome data. We treated exposure to co-twin or co-sibling violent victimisation as a time-varying covariate, with individuals considered unexposed until the first date of their co-relative having a recorded violent victimisation event. The nested data structure, with individuals being nested within multiple twin and sibling pairs, was accounted for using cluster-robust standard errors. We adjusted these models for the sex and birth year (continuous measure) of each proband and their co-relative. For monozygotic twins, we accounted for these confounders only in the proband, as they did not vary between co-twins. We pooled the estimates for the non-twin siblings available in both countries by fitting inverse variance weighted fixed-effects meta-analysis models.^25^

Under a genetic hypothesis, the magnitude of these associations would be expected to be largest among (a) monozygotic twins, who share all their co-segregating genes, followed by (b) dizygotic twins and non-twin full siblings, who share half on average, and (c) half-siblings, who share a quarter of their co-segregating genes on average. In contrast, a stronger association among maternal half-siblings compared to paternal half-siblings would indicate the presence of shared environmental influences, as maternal half-siblings share the same intrauterine environment and are more likely to experience greater shared environmental exposures due to the increased tendency for children to primarily reside with their mothers after parental separation.

### Quantitative Genetic Models

We fitted quantitative genetic structural equation models to estimate the relative contributions of additive genetic, shared environmental, and unique environmental factors (including measurement error and stochastic events) to individual differences in vulnerability to violent victimisation.^26^ Similar to previous work,^27,28^ we randomly selected one sibling pair from each family for these analyses. Genetic correlations between twins and siblings were fixed at their expected theoretical values: additive genetic correlations were set at 1.0 for monozygotic twins, 0.5 for dizygotic twins and non-twin full siblings, and 0.25 for half-siblings, reflecting the average proportion of co-segregating genes they share.

Shared environmental influences refer to all non-genetic factors that contribute to increased sibling similarities in violent victimisation risk. In the Swedish sample, where zygosity data were available, we examined shared environmental influences in two separate components. The first component represented environmental factors specific to twins, such as shared developmental stages and the timing of environmental exposures, with correlations fixed at 1.0 for twins and 0 for other siblings. The second component, included in models applied to both samples, encompassed shared environmental factors common to all siblings within a family. For this component, correlations were set at 1.0 for all sibling pairs, except for paternal half-siblings, where correlations were fixed at 0, as they were assumed to share a lower degree early-life environment. We comprehensively tested these assumptions (**eText 1**).

The quantitative genetic models were adjusted for birth year and sex. To account for the binary nature of the phenotype, liability-threshold models were specified, assuming that victimisation risk reflects an underlying normal distribution of liability, with a threshold distinguishing individuals with and without the observed phenotype.^29^ To account for differences in the rates of violent victimisation across sibling types, we relaxed the equal threshold assumption, which assumes a constant rate across all sibling types. The most parsimonious model in each country was selected on the basis of a combination of likelihood ratio tests and Akaike’s information criterion (AIC), and we subsequently pooled the variance components using the same meta-analytic approach as described above.

### Developmental periods

We conducted age-stratified analyses to examine how the relative contribution of genetic and environmental factors varied by age at first victimisation in three categories: childhood (0-11 years), adolescence (12-18 years), and adulthood (>18 years). We applied the same quantitative genetic modelling approach to each age group separately. For childhood analyses, models were estimated using Swedish data only, as the low number of childhood victimisation events in Finland (n=775; 0.04%) resulted in model non-convergence. For adolescence and adulthood analyses, we pooled data across both countries. Individuals were classified according to when they first experienced violence, with each individual contributing to only one developmental period analysis.

### Sex-limitation models

In separate analyses, we used sex-limitation models to test for quantitative and qualitative sex differences. Quantitative differences reflect variation in heritability magnitude between males and females, whilst qualitative differences reflect whether different genetic factors underlie victimisation risk across sexes. The latter are estimated by comparing same-sex and opposite-sex relatives. Qualitative differences are indicated when the genetic correlation between sexes deviates from unity (r_g_=1.0), evidenced by weaker phenotypic correlations in opposite-sex than same-sex relative pairs.

## Results

We examined a total of 5,296,997 individuals, of whom 154,209 (2.9%) had a recorded violent victimisation event throughout the follow-up period (**Table 1; eTable 1**). Males (4.0%) were over twice as likely to have had a recorded violent victimisation event compared to females (1.8%). Across both countries, we identified a total of 4,458,368 individuals (n_Sweden_=2,708,425; n_Finland_=1,749,943) who had at least one co-twin or co-sibling (**Table 2; eTable 2**). Compared to twins and non-twin full-siblings (2.2%-2.7%), we found that half-siblings had higher rates of violent victimisation (4.9%-5.4%) and more likely to have an immigrant background (**Table 2**).

**Table 1.** Descriptive characteristics of individuals included in the study pooled across Sweden and Finland.

|  | **Total sample**<br>**N (column**<br>**percent)** | **Exposed to violent**<br>**victimisation**<br>**N (row percent)** | **Unexposed to**<br>**violent**<br>**victimisation**<br>**N (row percent)** |
| --- | --- | --- | --- |
| *Sample* | 5,296,997<br>(100%) | 154,209 (2.9%) | 5,142,788 (97.1%) |
| *Sex* |  |  |  |
| Female | 2,580,214<br>(48.7%) | 46,164 (1.8%) | 2,534,050 (98.2%) |
| Male | 2,716,783<br>(51.3%) | 108,045 (4.0%) | 2,608,738 (96.0%) |
| *Immigrant*<br>*background* |  |  |  |
| No | 4,581,487<br>(86.5%) | 126,660 (2.8%) | 4,454,827 (97.2%) |
| Yes | 715,510 (13.5%) | 27,549 (3.9%) | 687,961 (96.1%) |
| *Birth years* |  |  |  |
| 1970-1974 | 509,893 (9.6%) | 12,089 (2.4%) | 497,804 (97.6%) |
| 1975-1979 | 800,117 (15.1%) | 21,050 (2.6%) | 779,067 (97.4%) |
| 1980-1984 | 784,851 (14.8%) | 26,121 (3.3%) | 758,730 (96.7%) |
| 1985-1989 | 832,604 (15.7%) | 34,080 (4.1%) | 798,524 (95.9%) |
| 1990-1994 | 915,746 (17.3%) | 33,162 (3.6%) | 882,584 (96.4%) |
| 1995-1999 | 755,708 (14.3%) | 18,645 (2.5%) | 737,063 (97.5%) |
| 2000-2004 | 698,078 (13.2%) | 9,062 (1.3%) | 689,016 (98.7%) |

**Table 2.** Descriptive characteristics of twins and siblings included in the study by relative type pooled across Sweden and Finland.

|  | **Total** | **Monozygotic twins** | **Dizygotic twins** | **Full siblings** | **Maternal half-siblings** | **Paternal half-siblings** |
| --- | --- | --- | --- | --- | --- | --- |
| *Unique individuals, n* | 4,455,431 | 14,954 | 28,010 | 3,950,781 | 629,323 | 659,800 |
| *Pairs used in familial aggregation analyses, n* | 8,839,097 | 14,954 | 28,010 | 6,505,700 | 960,644 | 1,058,957 |
| *Pairs used in quantitative genetic models, n* | 2,104,982 | 7,477 | 14,005 | 1,648,705 | 217,488 | 217,307 |
| *Male sex, n (%)* | 2,282,828 (51.2%) | 6,830 (45.7%) | 14,074 (50.2%) | 2,026,050 (51.3%) | 321,545 (51.1%) | 336,451 (51.0%) |
| *Immigrant background, n (%)* | 575,868 (12.9%) | 2,456 (16.4%) | 4,370 (15.6%) | 474,819 (12.0%) | 110,183 (17.5%) | 132,704 (20.1%) |
| *Birth years, n (%)* |  |  |  |  |  |  |
| 1970-1974 | 315,544 (7.1%) | 686 (4.6%) | 798 (2.8%) | 272,187 (6.9%) | 37,572 (6.0%) | 44,989 (6.8%) |
| 1975-1979 | 630,347 (14.1%) | 1,834 (12.3%) | 1,876 (6.7%) | 566,144 (14.3%) | 73,577 (11.7%) | 82,436 (12.5%) |
| 1980-1984 | 708,265 (15.9%) | 1,556 (10.4%) | 1,700 (6.1%) | 639,686 (16.2%) | 98,855 (15.7%) | 101,213 (15.3%) |
| 1985-1989 | 777,813 (17.5%) | 2,172 (14.5%) | 2,750 (9.8%) | 697,734 (17.7%) | 127,509 (20.3%) | 126,663 (19.2%) |
| 1990-1994 | 848,194 (19.0%) | 2,742 (18.3%) | 4,956 (17.7%) | 757,992 (19.2%) | 133,396 (21.2%) | 134,663 (20.4%) |
| 1995-1999 | 678,918 (15.2%) | 3,080 (20.6%) | 8,170 (29.2%) | 600,556 (15.2%) | 90,866 (14.4%) | 94,130 (14.3%) |
| 2000-2004 | 496,350 (11.1%) | 2,884 (19.3%) | 7,760 (27.7%) | 416,482 (10.5%) | 67,548 (10.7%) | 75,706 (11.5%) |
| *Violent victimisation* |  |  |  |  |  |  |
| Events, n (%) | 133,555 (3.0%) | 336 (2.2%) | 652 (2.3%) | 108,195 (2.7%) | 34,208 (5.4%) | 32,393 (4.9%) |
| Age at first event, mean years (SD) | 23.0 (7.0) | 22.5 (6.9) | 20.0 (5.7) | 23.1 (6.8) | 22.7 (7.0) | 22.8 (7.2) |

Individuals whose monozygotic co-twin had experienced violent victimisation were about six times as likely to experience violent victimisation themselves compared to those whose monozygotic co-twin was unaffected (adjusted hazard ratio, aHR=6.0; 95% CI: 4.0-9.0; **Figure 1; eTable 3**). Consistent with the presence of genetic influences, we found that this association was attenuated by approximately half among dizygotic twins and non-twin full siblings (aHR range: 2.4-2.6), who share on average 50% of their co-segregating genes, and was halved again among half-siblings (aHR range: 1.4-1.5), who share on average 25% of their co-segregating genes (**Figure 1**). The marginal differences in the associations between dizygotic twins and non-twin full-siblings suggest that the contributions of twin-specific environmental influences remain limited, and similarly, the marginal differences in the associations between maternal and paternal half-siblings suggest that the contributions of broader shared environmental influences are also limited.

**Figure 1.**
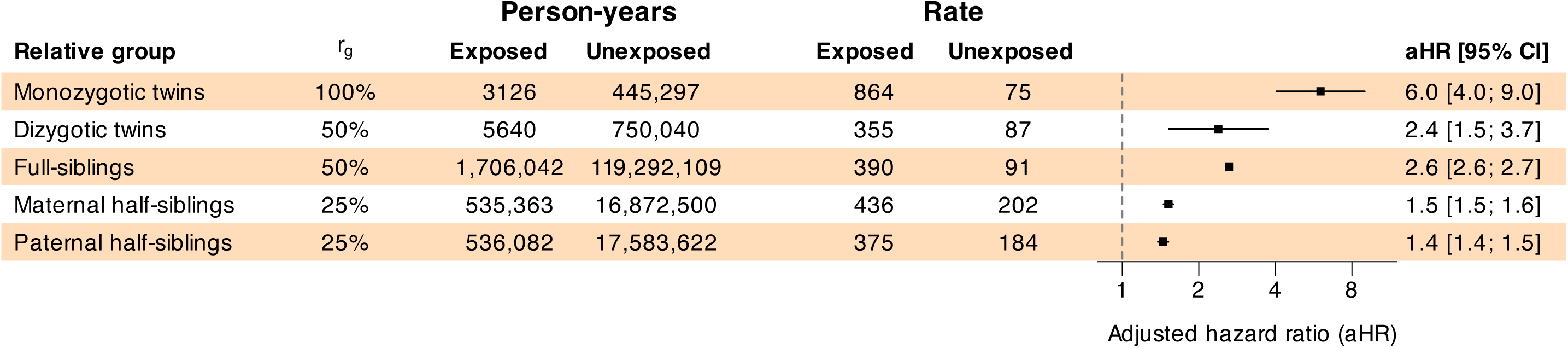
Familial aggregation of violent victimisation by degree of genetic relatedness pooled across Sweden and Finland. *Notes: r_g_, genetic correlation (average proportion of shared co-segregating genes); rate, the number of violent victimisation events per 100,000 person-years; aHR, adjusted hazard ratio, adjusted for the sex and birth year of the index person and the co-twin or co-sibling*.

The latter findings were confirmed in standard univariate quantitative genetic models, where the model fit indices (**eTable 4**) indicated that the best-fitting models included two components that captured additive genetic (i.e., heritability, h^2^) and unique environmental influences (e^2^). This implies that shared environmental influences (c^2^), including twin-specific environmental influences (t^2^), did not meaningfully explain individual differences in victimisation risk in either country, consistent with the correlational patterns observed in the familial aggregation analyses. In pooled cross-country analyses, additive genetic influences explained 47% (95% CI: 46%-49%; country-specific range: 45%-48%; **Figure 2**) of the variance in violent victimisation risk, and the remaining 53% (95% CI: 51%-54%) was attributed to unique environmental influences.

**Figure 2.**
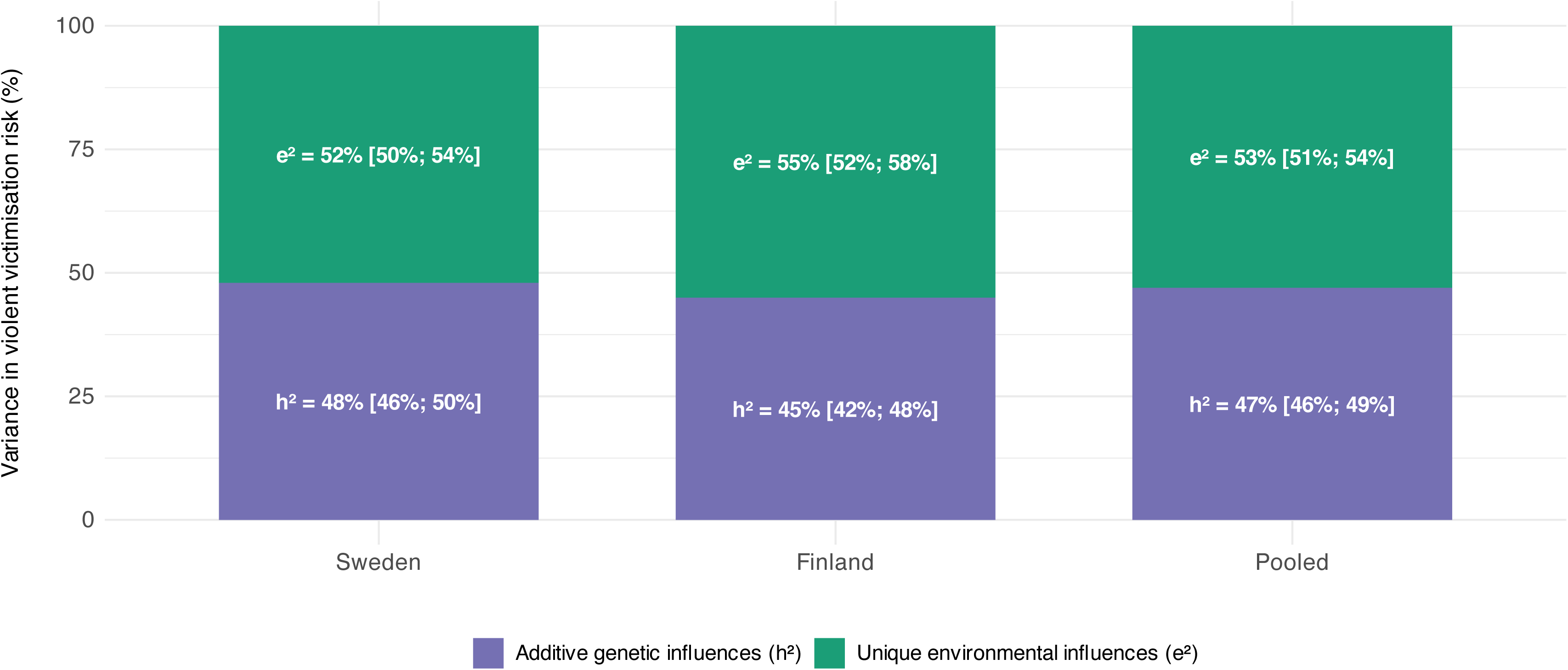
Decomposition of violent victimisation variance into additive genetic (h²) and unique environmental (e²) components for Sweden, Finland, and the pooled sample across both countries, according to the best-fitting models. Values represent percentages with 95% confidence intervals.

As a complementary sensitivity analysis, we analysed a broader violent victimisation phenotype using police-recorded data from Finland, which included 146,665 individuals (7.3% of the sample) with a similar average age at first victimisation as those ascertained using hospital and mortality records (24.8 vs. 25.5 years; p=0.741). We obtained similar results when we reran the quantitative genetic model on this alternative phenotype, although the shared environmental contribution to variance, albeit small, was statistically different from zero (h^2^=46%; 95% CI: 41%-52%; c^2^=5%; 95% CI: 3%-8%; e^2^=49%; 95% CI: 45%-51%). We further obtained consistent findings when we examined subsets of twins and non-twin siblings in Sweden (eFigure 1), assumed paternal half-siblings shared a larger proportion of their childhood environments, excluded right-censored individuals and those with an immigrant background, and analysed recurrent victimisation (h^2^: 42%-52%; e^2^: 48%-58%; eFigures 1-3).

In subgroup analyses, our findings were strongly moderated by the age at first victimisation event (**Figure 3; eTable 5**). Additive genetic factors accounted for a much larger proportion of the variance in violent victimisation risk during childhood compared to later developmental stages (h^2^: 70% vs. 40%-44%). Notably, shared environmental factors accounted for 22% (95% CI: 9%-35%) of the individual differences in violent victimisation risk during childhood, but none of the variance in subsequent developmental stages. The contributions of unique environmental influences on violent victimisation risk were not statistically significant during childhood (e^2^=8%; 95% CI: 0%-23%) but explained approximately 60% (e^2^ range: 56%-60%) of the individual differences observed during adolescence and adulthood.

**Figure 3.**
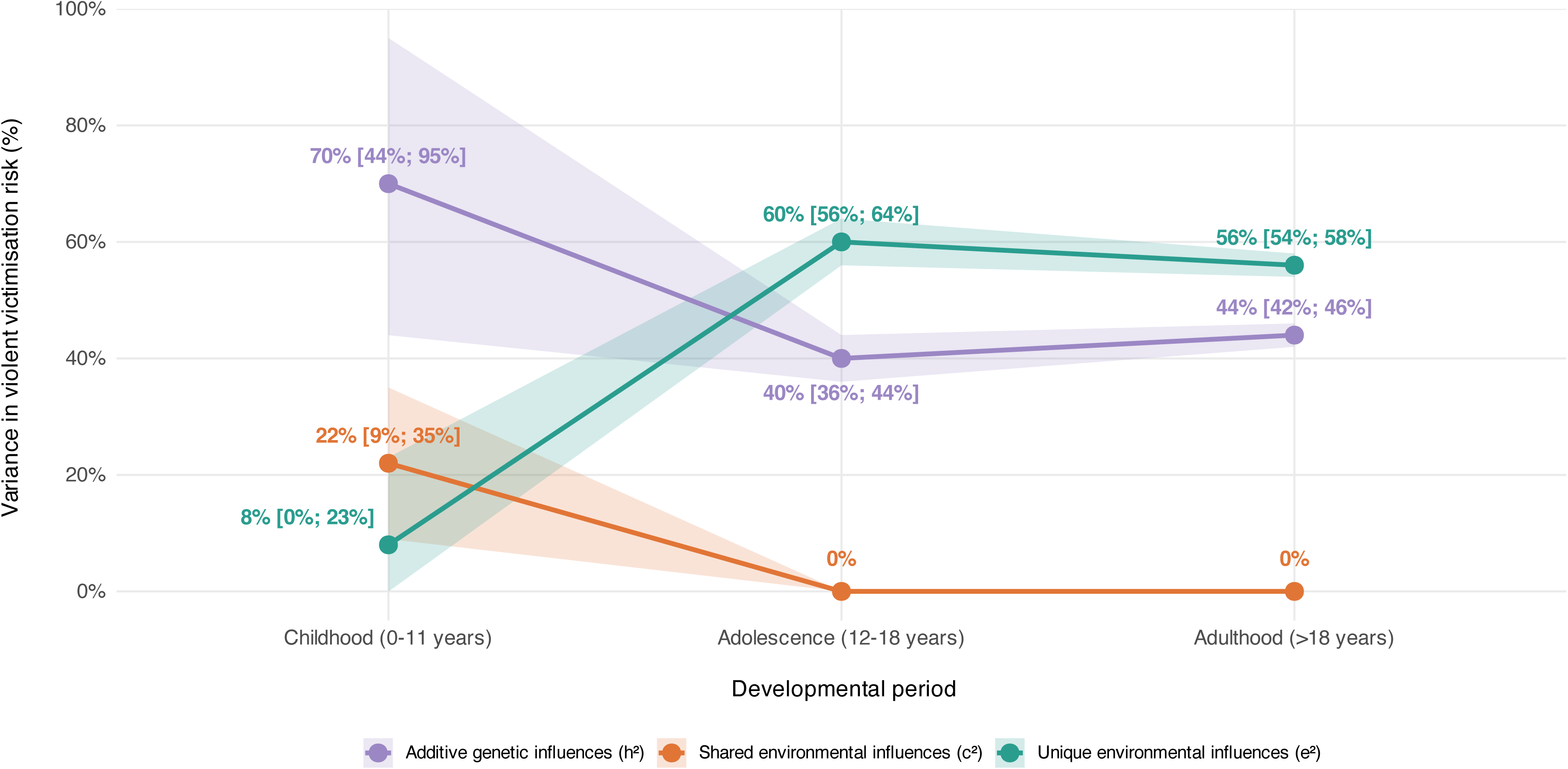
Decomposition of violent victimisation variance into additive genetic (h²), shared environmental (c²), and unique environmental (e²) components, stratified by age at first victimisation (childhood, adolescence, adulthood), pooled across both countries. Values represent percentages with 95% confidence intervals. For childhood analyses, only Swedish data were used due to the low number of Finnish incidents.

Sex-specific heritability estimates for violent victimisation demonstrated minimal quantitative differences, with additive genetic factors explaining between 52% and 54% of variance in females and males (**Figure 4A; eTables 6-8**). This negligible difference of 2% was not statistically significant (p=0.22). We replicated these findings using police reported victimisation events in Finland, where additive genetic influences accounted for 51% of the variance in both sexes (**Figure 4A**). In contrast, we found strong support for qualitative sex differences for both victimisation outcomes, meaning that partly distinct genetic factors contributed to the heritability of violent victimisation across sexes. The cross-sex genetic correlations (r_g_) ranged from 0.77 to 0.78, thus suggesting that approximately 60% (r_g_ ^2^) of the genetic liability was shared across sexes, whilst the remaining genetic variance was unique to each sex (**Figure 4B**).

**Figure 4.**
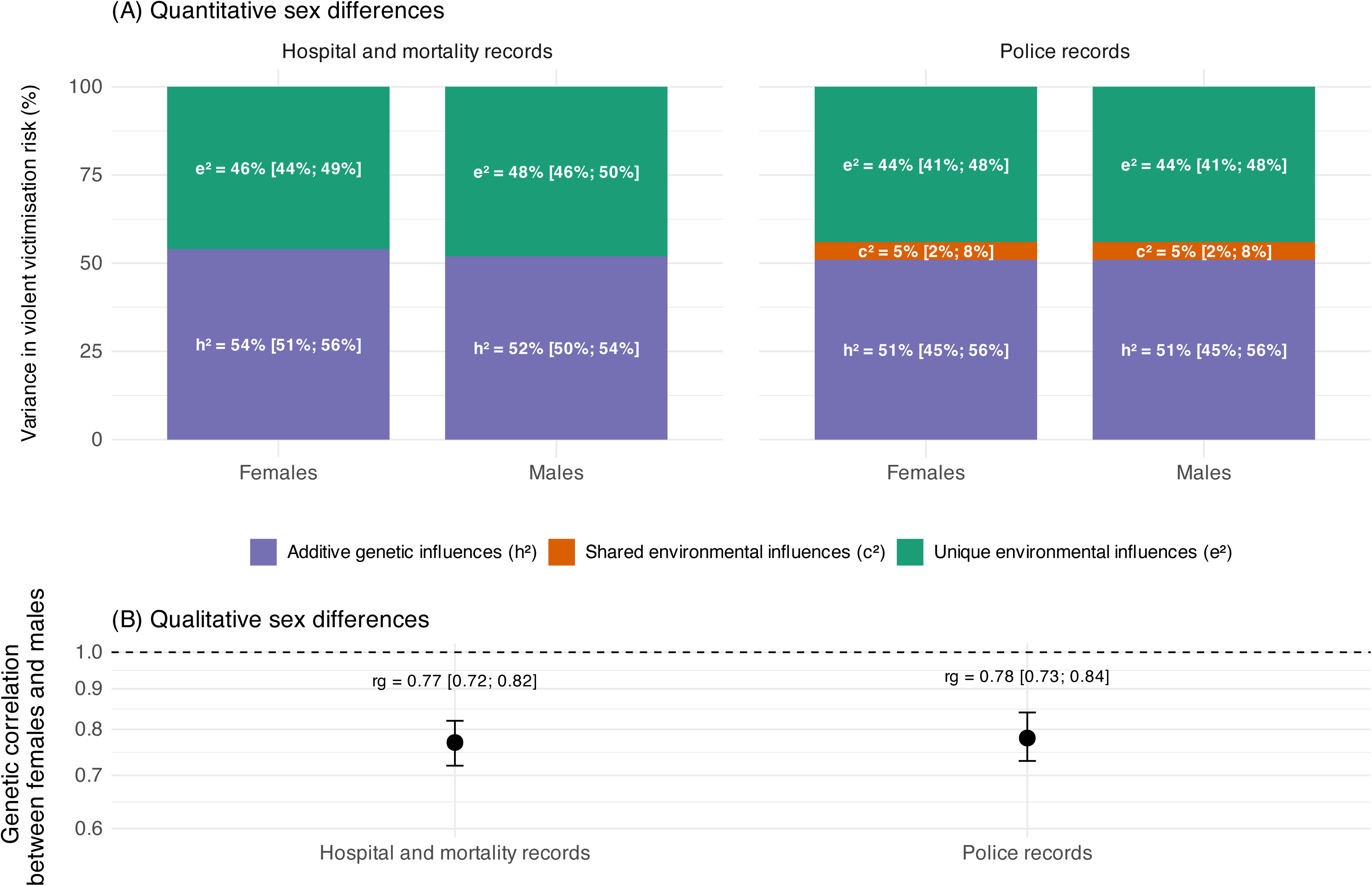
Quantitative and qualitative sex differences in genetic and environmental influences on violent victimisation. (A) Quantitative sex differences in the variance of violent victimisation risk decomposed into additive genetic (h²), shared environmental (c²), and unique environmental (e²) components, estimated separately for females and males using hospital and mortality records (left) and police records (right). Percentages and 95% confidence intervals are shown within bars. (B) Qualitative sex differences, expressed as genetic correlations (r_g_) between females and males, with 95% confidence intervals.

## Discussion

This cross-national study of 4.5 million Swedish and Finnish twins and siblings represents the largest investigation to date, to our knowledge, of genetic and environmental influences on violent victimisation risk. Our findings provide novel insights into the aetiology of violent victimisation across development and between sexes. We report two principal findings.

First, violent victimisation was strongly clustered within families, but the magnitude and sources of familial aggregation varied substantially across development. Familial aggregation analyses indicated that having a co-sibling who had been subjected to violence was associated with an approximately three-fold increase in one’s own risk. Consistent with the presence of genetic influences, the magnitude of this association increased with genetic relatedness, from approximately 1.5-fold among half-siblings to six-fold among monozygotic twins. When pooled across all ages, quantitative genetic models adjusting for sex and birth year indicated that 47% of the variance in violent victimisation risk was attributable to additive genetic factors. This estimate was replicated in Finland using police-reported victimisation (h^2^=46%) and proved robust to alternative sibling specifications, right-censoring, and potential confounding by immigrant background.

However, this overall estimate masks critical developmental heterogeneity in aetiological architecture. Childhood victimisation was predominantly explained by familial factors, with additive genetic effects explaining over two thirds (70%) of the variance and shared environmental factors accounting for an additional fifth (22%). By contrast, during adolescence and adulthood, this pattern shifted substantially, with additive genetic effects reducing to about 40%, shared environmental influences approaching zero, and unique environmental factors emerging as the dominant influence, accounting for the remaining variance.

Our findings show both consistencies and discrepancies with prior research. The overall heritability estimate (47%) is broadly consistent with previous family-based investigations into violent victimisation, which have typically reported heritability estimates in the range of 40-60%.^13–18^ Our developmental findings are only partly consistent with a recent systematic review^21^ of twin studies assessing broader indices of childhood victimisation, including maltreatment and bullying. Similar to the present findings, the review reported that shared environmental influences accounted for a moderate proportion of the variance in victimisation risk among younger children but for none of the corresponding variance among adolescents. In contrast to our findings, however, the review reported that additive genetic influences increased from 30% to 60% across these developmental stages, whereas we observed a decrease from 70% to approximately 40%.

These discrepancies likely reflect methodological differences between studies. More severe forms of violence requiring medical attention or police intervention may involve different aetiological pathways than milder victimisation experiences typically captured in parent- and self-report studies. Specifically, severe victimisation may be more strongly influenced by stable genetic vulnerabilities (such as those affecting emotional regulation or impulsivity) that persist across development, whereas milder forms may be more susceptible to age-related changes in social environments and peer influences that could alter the relative contributions of genetic and environmental factors. Childhood and adolescent/adult victimisation may also differ qualitatively, as childhood victimisation may be more characterised by parental abuse compared to adolescent/adult victimisation involving peer or stranger violence, which could have different aetiologies rather than simply reflecting age-related changes in the same underlying vulnerability. The considerable shared environmental influences observed in childhood are consistent with this interpretation, as young children typically share the same household environment as potentially abusive caregivers, while adolescents and adults experience more independent and diverse environmental exposures.

Beyond these substantive differences, measurement approaches may further contribute to discrepancies across studies. Our use of objective hospital and mortality records minimises reporter biases that have confounded earlier studies. Twin studies involving younger children rely more heavily on parental reports of victimisation events, whilst those on adolescents tend to focus on self-reports, making it impossible to disentangle temporal differences in aetiological influences from reporter biases.^22^ Future studies should test for the aetiological overlap between self-reported, parent-reported, and objective measures of victimisation, as well as between different types of victimisation (e.g., parental abuse, peer or stranger violence, and sexual victimisation), to determine whether these represent distinct phenotypes with different genetic architectures or variations of the same underlying liability.

Second, additive genetic influences demonstrated similar magnitudes in females and males, indicating minimal quantitative sex differences. However, comparisons of same-sex and opposite-sex pairs revealed a previously undetected pattern: genetic influences were only partly shared across sexes, with genetic correlations of 0.77-0.78 suggesting that approximately 40% of the heritable liability is sex-specific. This indicates that partly different constellations of genetic and environmental factors shape risk in males and females. This represents the first robust evidence of qualitative sex differences in the genetic architecture of violent victimisation, replicated across both countries, whereas only two previous studies^16,23^ have examined this phenomenon, and both appear to have lacked sufficient statistical power to detect such differences.

Relevant to interpreting these findings is that heritability estimates are probabilistic rather than deterministic,^30^ and likely operate indirectly through behavioural and social pathways. Genetic factors increase exposure to violence via parents’ partly heritable traits shaping the child’s environment (passive gene-environment correlation, rGE), through victims’ own partly heritable traits (e.g., neurodevelopmental and psychiatric disorders)^7,9,31^ eliciting responses from perpetrators (evocative rGE), and through partly genetically influenced selection into high-crime environments (active rGE). The sex-specific genetic effects suggest these pathways may operate differently in males and females through distinct risk factors or differential expression of genetic variants. Importantly, heritability estimates describe population-level patterns and should not be viewed as implying individual-level blame or inevitability.

Our findings have broader implications for future research by demonstrating the importance of distinguishing two distinct gene-environment processes. Previous studies of victimisation have rarely addressed genetic influences on trauma exposure itself (rGE), instead focusing on how genes moderate responses to traumatic events (gene-environment interaction, GxE).^32^ We have demonstrated that victimisation risk is non-randomly distributed and partly heritable, with genetic factors likely influencing both exposure to violent environments and characteristics that influence interactions with potential perpetrators. This has important methodological implications, as studies examining environmental effects on victimisation and its consequences must use family-based designs to account for unmeasured genetic confounding.^33–35^ Otherwise, putative environmental associations may instead reflect underlying genetic influences.

Whilst quantitative genetic models offer a powerful approach to estimating the heritability of violent victimisation, they cannot identify specific genetic variants, making genomic validation an important next step. Large-scale genotyped biobank cohorts linked to electronic health records would enable identification of individuals treated for assault-related injuries and examination of genetic variant associations. Our findings suggest that genome-wide association studies (GWAS) should be both age-stratified and sex-stratified. The developmental heterogeneity we identified indicates that pooling across ages would obscure distinct genetic architectures operating in childhood versus adolescence and adulthood. Similarly, the qualitative sex differences we observed suggest that pooling across sexes would likely obscure sex-specific genetic effects and the distinct pathways through which they operate. This also implies that downstream analyses, such as polygenic risk scores or Mendelian randomisation studies, should account for both developmental stage and sex-specific effects.

The study strengths included the use of nationwide register data that allowed us to study 32 entire birth cohorts from Sweden and Finland and their co-twins and siblings, which totalled more than 4.5 million individuals with negligible selection bias. Furthermore, by examining associations across multiple twin and sibling types within the same samples, we were able to explore the magnitude and nature of potential shared environmental influences in greater depth than earlier investigations. Additionally, the findings were replicated using two distinct objective measures of violent victimisation.

Two key limitations should be noted. First, whilst our approach of measuring violent victimisation using objective hospital and mortality records reduces reporting bias, it focuses on severe victimisation events requiring medical attention or resulting in death. However, our pooled heritability estimate (47%) remains consistent with earlier studies using parent- and self-reported measures that captured broader victimisation experiences (typically 40-60%), and we obtained similar results using police-recorded data in Finland (46%). This suggests that the genetic and environmental architecture of victimisation liability may be similar across different measurement approaches and severity levels, and their aetiological overlap could be investigated using multivariate quantitative genetic approaches in future studies.^33,36^ Second, the quantitative genetic models rest on several assumptions. Regarding the equal environments assumption for twins, our sensitivity analyses in the Swedish sample excluding twins entirely (eFigure 1) did not alter the heritability estimate (h^2^=0.48), indicating that potential violations of this assumption do not materially affect our conclusions. The relative proportion of heritability explained by dominance genetic effects remains minimal for most human traits,^37^ and simulation studies find that explicit modelling of such effects rarely alters the overall proportion of variance attributed to genetic factors.^38^ While non-random mating has been established for many phenotypes, we are unaware of studies estimating spousal correlations in violent victimisation measures,^39,40^ though accounting for direct non-random mating did not materially alter heritability of violent crime convictions in a similar Swedish pedigree study.^41^ More importantly, the familial aggregation analyses, which are independent of these particular modelling assumptions, were consistent with the quantitative genetic models.

The generalisability of our findings is supported by cross-national replication and similar self-reported violent victimisation rates to the global average (2.2%-3.5% versus 3.1%) in the most recent international comparison.^1^ However, both Sweden and Finland are welfare states with universal healthcare, which may limit applicability to countries with different social and health policies. Nevertheless, consistency with previous studies from diverse populations in high-income countries suggests the core aetiological mechanisms may be broadly generalisable to such contexts, but further replication, particularly in low- and middle-income countries, remains necessary.

## Conclusions

This investigation of over 4.5 million Swedish and Finnish twins and siblings represents the largest examination of genetic and environmental influences on violent victimisation to date. Our findings reveal that victimisation liability is moderately to considerably heritable, exhibits distinct aetiological patterns across developmental stages, and demonstrates qualitative sex differences in underlying genetic architecture. Our findings on the shifting balance of familial versus individual risk factors across development may potentially inform the timing and targeting of prevention efforts, though the efficacy of specific interventions requires additional empirical evaluation. These results emphasise the critical importance of genetically informative designs for disentangling genetic and environmental contributions and inform future molecular genetic investigations to identify specific genomic variants contributing to victimisation risk.

## Data Availability

Data may be obtained from a third party and are not publicly available. Due to Swedish and Finnish privacy legislation, individual-level data cannot be shared publicly. Researchers wishing to replicate this work can apply for data access through Statistics Sweden (https://www.scb.se/en/services/ordering-data-and-statistics/ordering-microdata/), the Swedish National Board of Health and Welfare (https://www.socialstyrelsen.se/en/statistics-and-data/registers/), and Findata (https://findata.fi/en/).

## Acknowledgements

AS and SF were supported by the NIHR Oxford Health Biomedical Research Centre (grant BRC-1215-20005). PM was supported by the European Research Council under the European Union’s Horizon 2020 research and innovation programme (grant agreement No 101019329), the Strategic Research Council (SRC) within the Research Council of Finland grants for ACElife (#352543-352572) and LIFECON (#345219), the Research Council of Finland profiling grant for SWAN (#136528219) and FooDrug (#136528212), and grants to the Max Planck – University of Helsinki Center from the Jane and Aatos Erkko Foundation (#210046), the Max Planck Society (#5714240218), University of Helsinki (#77204227), and Cities of Helsinki, Vantaa and Espoo (#4706914). MA was also supported by ACElife (#352543-352574).

