## Supplementary Materials for "Genetic and environmental architecture of violent victimisation across development and sex: A study of 4.5 million Nordic twins and siblings"

**eText 1.** Robustness checks

We conducted several complementary sensitivity analyses to assess the robustness of our findings. First, to validate the quantitative genetic model in Finland, which was based only on non-twin siblings, we refitted the Swedish model by excluding the twins to see whether the inclusion of twin data meaningfully altered the findings. We then tested assumptions about shared environmental correlations amongst half-siblings using two approaches. In the first, we reran the Swedish model including only twins to examine the impact of incorporating non-twin siblings with the standard half-sibling assumptions. In the second, we reran the models using all relative types in both countries but assuming that paternal half-siblings shared all childhood environments to obtain an upper bound of the extent to which this assumption influenced the estimates of the main model.

We also examined the effect of right-censoring by excluding individuals who had either died or emigrated during follow-up and assessed potential confounding by excluding those with an immigrant background. Finally, we identified individuals who had experienced violent victimisation on at least two separate occasions in Sweden (n=14,628; 0.5%) and Finland (n=5756; 0.3%). To minimise the risk of capturing follow-up care for the same incident, we required a minimum interval of 30 days between the first and second event. We then refitted the quantitative genetic models in both countries using this binary outcome, classifying individuals with at least two victimisation events as 1 and all others as 0.

**eTable 1. Descriptive characteristics of individuals included in the study by country**

|  | **Sweden** | | | **Finland** | | |
| --- | --- | --- | --- | --- | --- | --- |
|  | **Total sample  *N* (column percent)** | **Exposed to violent victimisation**  ***N* (row percent)** | **Unexposed to violent victimisation**  ***N* (row percent)** | **Total sample  *N* (column percent)** | **Exposed to violent victimisation**  ***N* (row percent)** | **Unexposed to violent victimisation**  ***N* (row percent)** |
| *Sample* | 3,227,205 (100%) | 109,398 (3.4%) | 3,117,807 (96.6%) | 2,069,792 (100%) | 44,811 (2.2%) | 2,024,981 (97.8%) |
| *Sex* |  |  |  |  |  |  |
| Female | 1,568,446 (48.6%) | 34,419 (2.2%) | 1,534,027 (97.8%) | 1,011,768 (48.9%) | 11,745 (1.2%) | 1,000,023 (98.8%) |
| Male | 1,658,759 (51.4%) | 74,979 (4.5%) | 1,583,780 (95.5%) | 1,058,024 (51.1%) | 33,066 (3.1%) | 1,024,958 (96.9%) |
| *Immigrant background* |  |  |  |  |  |  |
| No | 2,587,395 (80.2%) | 83,487 (3.2%) | 2,503,908 (96.8%) | 1,994,092 (96.3%) | 43,173 (2.2%) | 1,950,919 (97.8%) |
| Yes | 639,810 (19.8%) | 25,911 (4.0%) | 613,899 (96.0%) | 75,700 (3.7%) | 1638 (2.2%) | 74,062 (97.8%) |
| *Birth years* |  |  |  |  |  |  |
| 1970-1974 | 217,071 (6.7%) | 5268 (2.4%) | 211,803 (97.6%) | 292,822 (14.1%) | 6821 (2.3%) | 285,001 (97.7%) |
| 1975-1979 | 482,403 (14.9%) | 13,583 (2.8%) | 468,820 (97.2%) | 317,714 (15.4%) | 7467 (2.4%) | 310,247 (97.6%) |
| 1980-1984 | 465,499 (14.4%) | 18,049 (3.9%) | 447,450 (96.1%) | 319,352 (15.4%) | 8072 (2.5%) | 311,280 (97.5%) |
| 1985-1989 | 528,469 (16.4%) | 26,308 (5.0%) | 502,161 (95.0%) | 304,135 (14.7%) | 7772 (2.6%) | 296,363 (97.4%) |
| 1990-1994 | 593,806 (18.4%) | 25,920 (4.4%) | 567,886 (95.6%) | 321,940 (15.6%) | 7242 (2.2%) | 314,698 (97.8%) |
| 1995-1999 | 462,982 (14.3%) | 13,374 (2.9%) | 449,608 (97.1%) | 292,726 (14.1%) | 5271 (1.8%) | 287,455 (98.2%) |
| 2000-2004 | 476,975 (14.8%) | 6896 (1.4%) | 470,079 (98.6%) | 221,103 (10.7%) | 2166 (1.0%) | 218,937 (99.0%) |

**eTable 2. Descriptive characteristics of twins and siblings included in the study by country and relative type**

|  | **Total** | **Monozygotic twins** | **Dizygotic twins** | **Full siblings** | **Maternal half-siblings** | **Paternal half-siblings** |
| --- | --- | --- | --- | --- | --- | --- |
| ***Sweden*** |  |  |  |  |  |  |
| *Unique individuals, n* | 2,708,728 | 14,954 | 28,010 | 2,359,635 | 419,672 | 439,609 |
| *Pairs used in familial aggregation analyses, n* | 5,182,628 | 14,954 | 28,010 | 3,595,255 | 642,491 | 709,387 |
| *Pairs used in quantitative genetic models, n* | 1,320,285 | 7477 | 14,005 | 1,008,187 | 145,597 | 145,019 |
| *Male sex, n (%)* | 1,391,764 (51.4%) | 6830 (45.7%) | 14,074 (50.2%) | 1,214,362 (51.5%) | 214,955 (51.2%) | 224,898 (51.2%) |
| *Immigrant background, n (%)* | 515,495 (19.0%) | 2456 (16.4%) | 4370 (15.6%) | 424,685 (18.0%) | 100,166 (23.9%) | 117,629 (26.8%) |
| *Birth years, n (%)* |  |  |  |  |  |  |
| 1970-1974 | 123,307 (4.6%) | 686 (4.6%) | 798 (2.8%) | 99,109 (4.2%) | 19,327 (4.6%) | 22,631 (5.1%) |
| 1975-1979 | 364,657 (13.5%) | 1834 (12.3%) | 1876 (6.7%) | 320,432 (13.6%) | 48,241 (11.5%) | 53,682 (12.2%) |
| 1980-1984 | 418,278 (15.4%) | 1556 (10.4%) | 1700 (6.1%) | 372,511 (15.8%) | 64,398 (15.3%) | 65,646 (14.9%) |
| 1985-1989 | 494,707 (18.3%) | 2172 (14.5%) | 2750 (9.8%) | 438,717 (18.6%) | 88,476 (21.1%) | 87,920 (20.0%) |
| 1990-1994 | 549,701 (20.3%) | 2742 (18.3%) | 4956 (17.7%) | 485,366 (20.6%) | 92,669 (22.1%) | 94,452 (21.5%) |
| 1995-1999 | 418,412 (15.4%) | 3080 (20.6%) | 8170 (29.2%) | 363,159 (15.4%) | 59,271 (14.1%) | 61,760 (14.0%) |
| 2000-2004 | 339,666 (12.5%) | 2884 (19.3%) | 7760 (27.7%) | 280,341 (11.9%) | 47,290 (11.3%) | 53,518 (12.2%) |
| *Violent victimisation* |  |  |  |  |  |  |
| Events, n (%) | 95,352 (3.5%) | 336 (2.2%) | 652 (2.3%) | 75,824 (3.2%) | 25,873 (6.2%) | 24,696 (5.6%) |
| Age at first event,  mean years (SD) | 22.1 (6.4) | 22.5 (6.9) | 20.0 (5.7) | 22.1 (6.2) | 22.0 (6.5) | 22.1 (6.6) |
| ***Finland*** |  |  |  |  |  |  |
| *Unique individuals, n* | 1,746,703 | N/A | N/A | 1,591,146 | 209,651 | 220,191 |
| *Pairs used in familial aggregation analyses, n* | 3,656,469 | N/A | N/A | 2,910,445 | 318,153 | 349,570 |
| *Pairs used in quantitative genetic models, n* | 784,697 | N/A | N/A | 640,518 | 71,891 | 72,288 |
| *Male sex, n (%)* | 891,064 (51.0%) | N/A | N/A | 811,688 (51.0%) | 106,590 (50.8%) | 111,553 (50.7%) |
| *Immigrant background, n (%)* | 60,373 (3.5%) | N/A | N/A | 50,134 (3.2%) | 10,017 (4.8%) | 15,075 (6.8%) |
| *Birth years, n (%)* |  |  |  |  |  |  |
| 1970-1974 | 192,237 (11.0%) | N/A | N/A | 173,078 (10.9%) | 18,245 (8.7%) | 22,358 (10.2%) |
| 1975-1979 | 265,690 (15.2%) | N/A | N/A | 245,712 (15.4%) | 25,336 (12.1%) | 28,754 (13.1%) |
| 1980-1984 | 289,987 (16.6%) | N/A | N/A | 267,175 (16.8%) | 34,457 (16.4%) | 35,567 (16.2%) |
| 1985-1989 | 283,106 (16.2%) | N/A | N/A | 259,017 (16.3%) | 39,033 (18.6%) | 38,743 (17.6%) |
| 1990-1994 | 298,493 (17.1%) | N/A | N/A | 272,626 (17.1%) | 40,727 (19.4%) | 40,211 (18.3%) |
| 1995-1999 | 260,506 (14.9%) | N/A | N/A | 237,397 (14.9%) | 31,595 (15.1%) | 32,370 (14.7%) |
| 2000-2004 | 156,684 (9.0%) | N/A | N/A | 136,141 (8.6%) | 20,258 (9.7%) | 22,188 (10.1%) |
| *Violent victimisation* |  |  |  |  |  |  |
| Events, n (%) | 38,203 (2.2%) | N/A | N/A | 32,371 (2.0%) | 8335 (4.0%) | 7697 (3.5%) |
| Age at first event,  mean years (SD) | 25.4 (7.9) | N/A | N/A | 25.5 (7.7) | 24.9 (8.1) | 24.9 (8.4) |

**eTable 3. Familial aggregation of violent victimisation by country and degree of genetic relatedness**

|  | **r_g_** | **Person-years at risk** | | **Rate per 100,000 person-years** | | **aHR [95% CI]** |
| --- | --- | --- | --- | --- | --- | --- |
|  |  | **Exposed** | **Unexposed** | **Exposed** | **Unexposed** |  |
| ***Sweden*** |  |  |  |  |  |  |
| Monozygotic twins | 100% | 3126 | 445,297 | 864 | 75 | 6.0 [4.0; 9.0] |
| Dizygotic twins | 50% | 5640 | 750,040 | 355 | 87 | 2.4 [1.5; 3.7] |
| Full-siblings | 50% | 1,216,963 | 69,485,136 | 416 | 109 | 2.5 [2.5; 2.6] |
| Maternal half-siblings | 25% | 418,592 | 11,185,215 | 462 | 231 | 1.5 [1.4; 1.6] |
| Paternal half-siblings | 25% | 423,941 | 11,603,416 | 401 | 212 | 1.5 [1.4; 1.5] |
| ***Finland*** |  |  |  |  |  |  |
| Full-siblings | 50% | 489,079 | 49,836,070 | 324 | 65 | 3.0 [2.8; 3.2] |
| Maternal half-siblings | 25% | 116,771 | 5,702,834 | 345 | 146 | 1.5 [1.4; 1.7] |
| Paternal half-siblings | 25% | 112,141 | 6,008,599 | 276 | 129 | 1.4 [1.2; 1.6] |

*Notes: r_g_, genetic correlation (average proportion of shared co-segregating genes), aHR, adjusted hazard ratio, adjusted for the sex and birth year of the index person and the co-twin or co-sibling.*

**eTable 4. Variance component estimates and model fit statistics for violent victimisation by country**

|  | **EP** | **-2LL** | **df** | **AIC** | **p** | **h^2^** | **t^2^** | **c^2^** | **e^2^** |
| --- | --- | --- | --- | --- | --- | --- | --- | --- | --- |
| ***Sweden*** |  |  |  |  |  |  |  |  |  |
| ATCE | 11 | 831**,**456 | 2,640,560 | 831,478 | - | 0.46 [0.40; 0.51] | 0.03 [0.00; 0.09] | 0.01 [0.00; 0.04] | 0.50 [0.44; 0.57] |
| ATE | 10 | 831**,**457 | 2,640,561 | 831,477 | 0.404 | 0.48 [0.46; 0.50] | 0.02 [0.00; 0.09] | 0 (fixed) | 0.50 [0.43; 0.56] |
| ACE | 10 | 831**,**457 | 2,640,561 | 831,477 | 0.416 | 0.46 [0.41; 0.52] | 0 (fixed) | 0.01 [0.00; 0.04] | 0.53 [0.50; 0.56] |
| **AE** | **9** | **831,457** | **2**,**640**,**562** | **831**,**475** | **0.566** | **0.48 [0.46; 0.50]** | **0 (fixed)** | **0 (fixed)** | **0.52 [0.50; 0.54]** |
| ***Finland*** |  |  |  |  |  |  |  |  |  |
| ACE | 8 | 341**,**387 | 1,569,387 | 341,403 | - | 0.36 [0.25; 0.47] | N/A | 0.05 [0.00; 0.10] | 0.60 [0.54; 0.66] |
| **AE** | **7** | **341,390** | **1**,**569**,**388** | **341**,**404** | **0.082** | **0.45 [0.42; 0.48]** | **N/A** | **0 (fixed)** | **0.55 [0.52; 0.58]** |

*Notes: EP, estimated parameters. AIC, Akaike’s Information Criterion. P, p-value for likelihood ratio test against the ‘ACE’ model, h^2^, heritability, t^2^, twin-specific environmental influences, c^2^, common/shared environmental influences, e^2^, unique environmental influences. Model in bold represents the most parsimonious model on the basis of AIC. Lower bounds of confidence intervals were constrained at zero, as variance components cannot be negative.*

**eTable 5. Variance component estimates and model fit statistics for violent victimisation by country and age at first victimisation**

|  |  | **EP** | **-2LL** | **df** | **AIC** | **p** | **h^2^** | **c^2^** | **e^2^** |
| --- | --- | --- | --- | --- | --- | --- | --- | --- | --- |
| ***Sweden*** |  |  |  |  |  |  |  |  |  |
|  | ***Childhood*** |  |  |  |  |  |  |  |  |
|  | **ACE** | **10** | **39,052** | **2**,**640**,**561** | **39**,**072** | **-** | **0.70 [0.44; 0.95]** | **0.22 [0.09; 0.35]** | **0.08 [0.00; 0.23]** |
|  | AE | 9 | 39,071 | 2,640,562 | 39,089 | <0.001 | 0.99 [0.96; 1.01] | 0 (fixed) | 0.01 [0.00; 0.04] |
|  | ***Adolescence*** |  |  |  |  |  |  |  |  |
|  | ACE | 10 | 287,493 | 2,640,561 | 287,513 | - | 0.40 [0.36; 0.44] | 0.00 [0.00; 0.00] | 0.60 [0.56; 0.64] |
|  | **AE** | **9** | **287,493** | **2**,**640**,**562** | **287**,**511** | **1.000** | **0.40 [0.36; 0.44]** | **0** (fixed) | **0.60 [0.56; 0.64]** |
|  | ***Adulthood*** |  |  |  |  |  |  |  |  |
|  | ACE | 10 | 633,714 | 2,640,561 | 633,734 | - | 0.41 [0.34; 0.48] | 0.01 [0.00; 0.05] | 0.58 [0.54; 0.62] |
|  | **AE** | **9** | **633**,**715** | **2**,**640**,**562** | **633**,**733** | **0.384** | **0.44 [0.42; 0.46]** | **0 (fixed)** | **0.56 [0.54; 0.58]** |
| ***Finland*** |  |  |  |  |  |  |  |  |  |
|  | ***Adolescence*** |  |  |  |  |  |  |  |  |
|  | ACE | 8 | 71,199 | 1,569,387 | 71,215 | - | 0.22 [0.00; 0.60] | 0.10 [0.00; 0.27] | 0.68 [0.47; 0.89] |
|  | **AE** | **7** | **71,200** | **1,569,388** | **71,214** | **0.279** | 0.42 [0.36; 0.44] | **0 (fixed)** | 0.58 [0.48; 0.68] |
|  | ***Adulthood*** |  |  |  |  |  |  |  |  |
|  | ACE | 8 | 294,721 | 1,569,387 | 294,737 | - | 0.36 [0.24; 0.49] | 0.03 [0.00; 0.09] | 0.61 [0.54; 0.68] |
|  | **AE** | **7** | **294,722** | **1,569,388** | **294,736** | **0.381** | **0.42 [0.39; 0.45]** | **0 (fixed)** | **0.58 [0.55; 0.61]** |

*Notes: EP, estimated parameters. AIC, Akaike’s Information Criterion. P, p-value for likelihood ratio test against the ‘ACE’ model, h^2^, heritability, c^2^, common/shared environmental influences, e^2^, unique environmental influences. Model in bold represents the most parsimonious model on the basis of AIC. Lower bounds of confidence intervals were constrained at zero, as variance components cannot be negative. The childhood model was not sufficiently powered to be estimated in Finland.*

**eFigure 1. Sensitivity analyses in the Swedish sample comparing the main model with models excluding non-twin siblings, twins, right-censored individuals, and second-generation immigrants. Percentages with 95% CIs.**

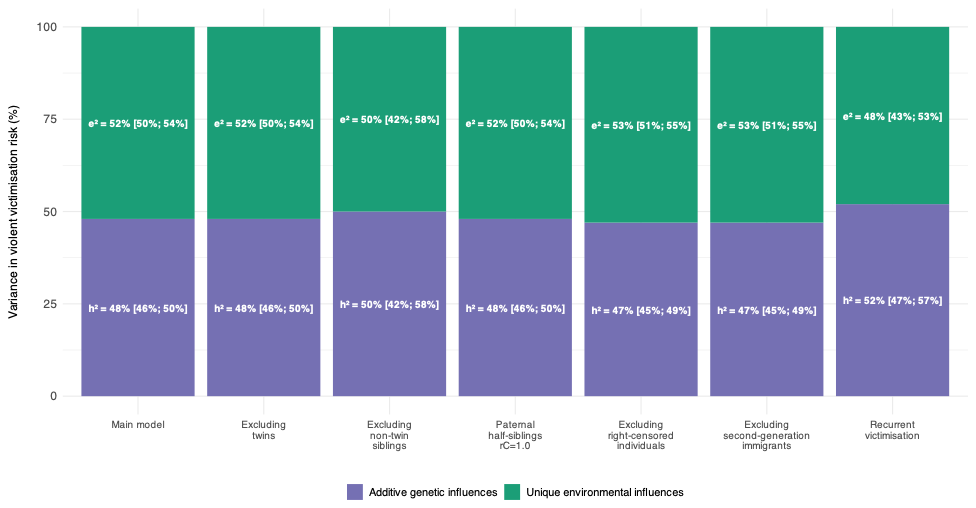

**eFigure 2. Sensitivity analyses in the Finnish sample comparing the main model with models excluding right-censored individuals and second-generation immigrants. Percentages with 95% CIs.**

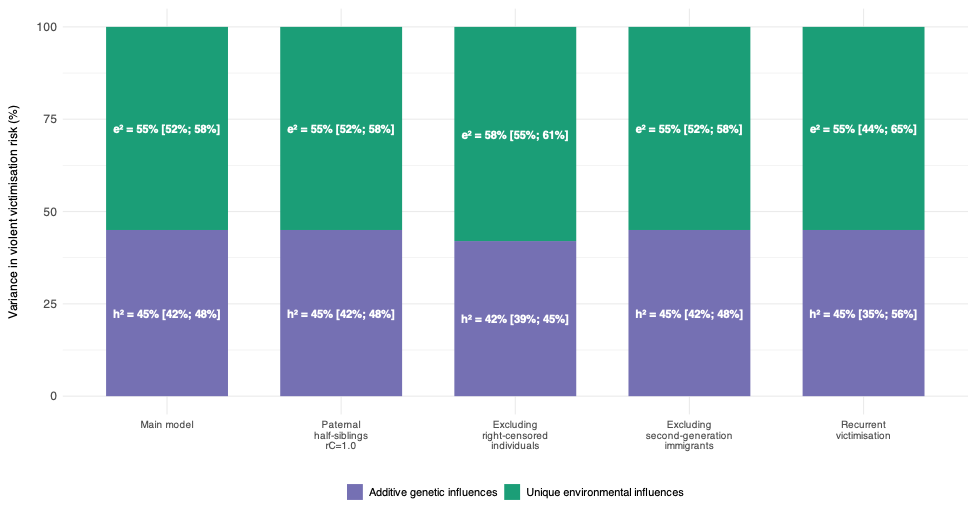

**eFigure 3. Sensitivity analyses in the pooled sample across Sweden and Finland comparing the main model with models excluding right-censored individuals and second-generation immigrants. Percentages with 95% CIs.**

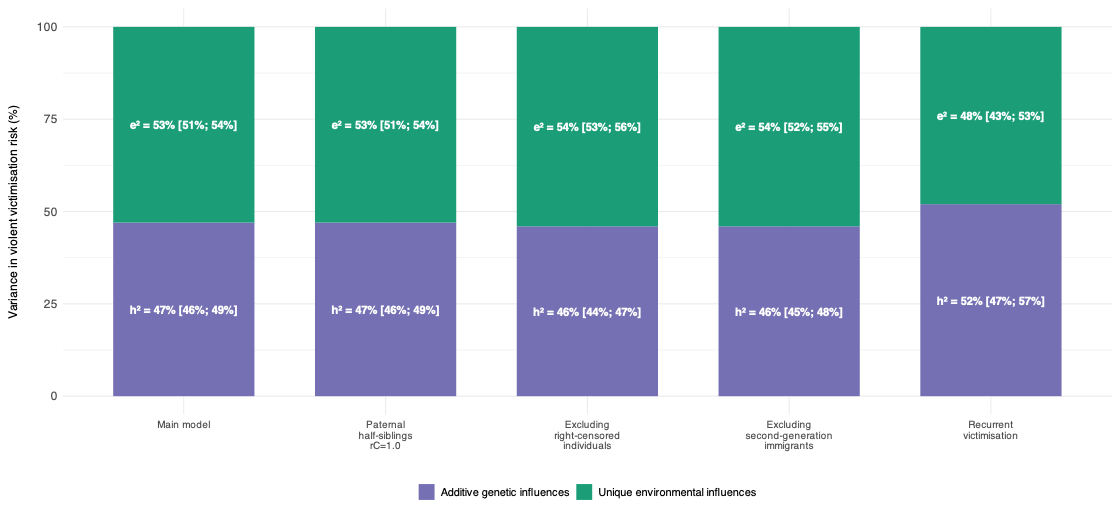

**eTable 6. Model fit statistics and parameter estimates from sex-limitation models of genetic and environmental variance components for violent victimisation in Sweden**

|  |  |  |  |  |  | **Females** | | | **Males** | | | **r_G_(sex)**  **[95% CI]** |
| --- | --- | --- | --- | --- | --- | --- | --- | --- | --- | --- | --- | --- |
| **Model** | **EP** | **-2LL** | **df** | **AIC** | ***P*** | **h^2^**  **[95% CI]** | **c^2^**  **[95% CI]** | **e^2^**  **[95% CI]** | **h^2^**  **[95% CI]** | **c^2^**  **[95% CI]** | **e^2^**  **[95% CI]** |  |
| Saturated | 43 | 831269.4 | 2640527 | 831355.4 | - | - | - | - | - | - | - | - |
| aceQuantQual | 24 | 831,286 | 2,640,546 | 831,334 | 0.590 | 0.52 [0.41; 0.62] | 0.02 [0.00; 0.07] | 0.47 [0.41; 0.53] | 0.50 [0.42; 0.58] | 0.01 [0.00; 0.05] | 0.49 [0.44; 0.53] | 0.77 [0.70; 0.84] |
| aceQual | 22 | 831,287 | 2,640,548 | 831,331 | 0.663 | 0.50 [0.45; 0.56] | 0.01 [0.00; 0.04] | 0.48 [0.45; 0.52] | 0.50 [0.45; 0.56] | 0.01 [0.00; 0.04] | 0.48 [0.45; 0.52] | 0.77 [0.71; 0.84] |
| aceQuant | 23 | 831,295 | 2,640,547 | 831,341 | 0.178 | 0.36 [0.26; 0.46] | 0.09 [0.04; 0.14] | 0.55 [0.49; 0.62] | 0.52 [0.49; 0.55] | 0.00 [0.00; 0.00] | 0.48 [0.45; 0.51] | 1 (fixed) |
| ace | 21 | 831,328 | 2,640,549 | 831,370 | <0.001 | 0.46 [0.41; 0.52] | 0.01 [0.00; 0.04] | 0.53 [0.50; 0.56] | 0.46 [0.41; 0.52] | 0.01 [0.00; 0.04] | 0.53 [0.50; 0.56] | 1 (fixed) |
| aeQuantQual | 22 | 831,288 | 2,640,548 | 831,332 | 0.642 | 0.55 [0.50; 0.59] | 0 (fixed) | 0.45 [0.41; 0.50] | 0.52 [0.50; 0.55] | 0 (fixed) | 0.48 [0.45; 0.50] | 0.78 [0.72; 0.84] |
| **aeQual** | **21** | **831**,**288** | **2**,**640**,**549** | **831**,**330** | **0.654** | **0.53 [0.51; 0.55]** | **0 (fixed)** | **0.47 [0.45; 0.49]** | **0.53 [0.51; 0.55]** | **0 (fixed)** | **0.47 [0.45; 0.49]** | **0.79 [0.73; 0.85]** |
| aeQuant | 21 | 831,327 | 2,640,549 | 831,369 | <0.001 | 0.46 [0.43; 0.50] | 0 (fixed) | 0.54 [0.50; 0.57] | 0.49 [0.47; 0.52] | 0 (fixed) | 0.51 [0.48; 0.53] | 1 (fixed) |
| ae | 20 | 831,328 | 2,640,550 | 831368 | <0.001 | 0.48 [0.46; 0.50] | 0 (fixed) | 0.52 [0.50; 0.54] | 0.48 [0.46; 0.50] | 0 (fixed) | 0.52 [0.50; 0.54] | 1 (fixed) |

*Notes: EP, estimated parameters. AIC, Akaike’s Information Criterion. P, p-value for likelihood ratio test against the saturated model, h^2^, heritability, c^2^, common/shared environmental influences, e^2^, unique environmental influences, rG(sex), cross-sex genetic correlation estimated using opposite-sexed sibling pairs, 95% CI, 95% confidence intervals. Saturated refers to the saturated model. aceQuantQual is a quantitative genetic ‘ACE’ model estimating additive genetic influences (A), common environmental influences (C) and unique environmental influences (E), allowing for both qualitative and quantitative sex differences. aceQual and aceQuant are ACE models that either allow for qualitative or quantitative sex differences, respectively, whilst ace refers to an ACE model without any sex differences. The AE-models, which are variants of ACE models that exclude common environmental influences, were named similarly. Model in bold represents the most parsimonious model on the basis of AIC. Lower bounds of confidence intervals were constrained at zero, as variance components cannot be negative.*

**eTable 7. Model fit statistics and parameter estimates from sex-limitation models of genetic and environmental variance components for violent victimisation in Finland**

|  |  |  |  |  |  | **Females** | | | **Males** | | | **r_G_(sex)**  **[95% CI]** |
| --- | --- | --- | --- | --- | --- | --- | --- | --- | --- | --- | --- | --- |
| **Model** | **EP** | **-2LL** | **df** | **AIC** | ***P*** | **h^2^**  **[95% CI]** | **c^2^**  **[95% CI]** | **e^2^**  **[95% CI]** | **h^2^**  **[95% CI]** | **c^2^**  **[95% CI]** | **e^2^**  **[95% CI]** |  |
| Saturated | 28 | 341,335 | 1,569,366 | 341,391 | - | - | - | - | - | - | - | - |
| aceQuantQual | 18 | 341,350 | 1,569,376 | 341,386 | 0.155 | 0.42 [0.04; 0.80] | 0.09 [0.00; 0.27] | 0.49 [0.29; 0.70] | 0.42 [0.23; 0.62] | 0.03 [0.00; 0.12] | 0.55 [0.44; 0.65] | 0.66 [0.36; 0.97] |
| aceQual | 16 | 341,356 | 1,569,378 | 341,388 | 0.055 | 0.41 [0.30; 0.53] | 0.05 [0.00; 0.10] | 0.54 [0.47; 0.61] | 0.41 [0.30; 0.53] | 0.05 [0.00; 0.10] | 0.54 [0.47; 0.61] | 0.69 [0.54; 0.84] |
| aceQuant | 17 | 341,351 | 1,569,377 | 341,385 | 0.166 | 0.28 [0.03; 0.54] | 0.15 [0.03; 0.27] | 0.56 [0.42; 0.71] | 0.48 [0.43; 0.53] | 0.00 [0.00; 0.01] | 0.52 [0.48; 0.57] | 1 (fixed) |
| ace | 15 | 341,373 | 1,569,379 | 341,403 | <0.001 | 0.36 [0.25; 0.47] | 0.05 [0.00; 0.10] | 0.60 [0.53; 0.66] | 0.36 [0.25; 0.47] | 0.05 [0.00; 0.10] | 0.60 [0.53; 0.66] | 1 (fixed) |
| **aeQuantQual** | **16** | **341**,**353** | **1**,**569**,**378** | **341**,**385** | **0.127** | **0.60 [0.52; 0.67]** | **0 (fixed)** | **0.40 [0.33; 0.48]** | **0.48 [0.43; 0.52]** | **0 (fixed)** | **0.52 [0.48; 0.57]** | **0.71 [0.60; 0.81]** |
| aeQual | 15 | 341,359 | 1,569,379 | 341,389 | 0.033 | 0.51 [0.47; 0.55] | 0 (fixed) | 0.49 [0.45; 0.53] | 0.51 [0.47; 0.55] | 0 (fixed) | 0.49 [0.45; 0.53] | 0.75 [0.64; 0.86] |
| aeQuant | 15 | 341,376 | 1,569,379 | 341,406 | <0.001 | 0.48 [0.41; 0.55] | 0 (fixed) | 0.52 [0.45; 0.59] | 0.44 [0.39; 0.48] | 0 (fixed) | 0.56 [0.52; 0.61] | 1 (fixed) |
| ae | 14 | 341,376 | 1,569,380 | 341,404 | <0.001 | 0.45 [0.42; 0.48] | 0 (fixed) | 0.55 [0.52; 0.58] | 0.45 [0.42; 0.48] | 0 (fixed) | 0.55 [0.52; 0.58] | 1 (fixed) |

*Notes: EP, estimated parameters. AIC, Akaike’s Information Criterion. P, p-value for likelihood ratio test against the saturated model, h^2^, heritability, c^2^, common/shared environmental influences, e^2^, unique environmental influences, rG(sex), cross-sex genetic correlation estimated using opposite-sexed sibling pairs, 95% CI, 95% confidence intervals. Saturated refers to the saturated model. aceQuantQual is a quantitative genetic ‘ACE’ model estimating additive genetic influences (A), common environmental influences (C) and unique environmental influences (E), allowing for both qualitative and quantitative sex differences. aceQual and aceQuant are ACE models that either allow for qualitative or quantitative sex differences, respectively, whilst ace refers to an ACE model without any sex differences. The AE-models, which are variants of ACE models that exclude common environmental influences, were named similarly. Model in bold represents the most parsimonious model on the basis of AIC. Lower bounds of confidence intervals were constrained at zero, as variance components cannot be negative.*

**eTable 8. Model fit statistics and parameter estimates from sex-limitation models of genetic and environmental variance components for police-reported violent victimisation in Finland**

|  |  |  |  |  |  | **Females** | | | **Males** | | | **r_G_(sex)**  **[95% CI]** |
| --- | --- | --- | --- | --- | --- | --- | --- | --- | --- | --- | --- | --- |
| **Model** | **EP** | **-2LL** | **df** | **AIC** | ***P*** | **h^2^**  **[95% CI]** | **c^2^**  **[95% CI]** | **e^2^**  **[95% CI]** | **h^2^**  **[95% CI]** | **c^2^**  **[95% CI]** | **e^2^**  **[95% CI]** |  |
| Saturated | 28 | 799,363 | 1,506,094 | 799,419 | - | - | - | - | - | - | - | - |
| aceQuantQual | 18 | 799,379 | 1,506,104 | 799,415 | 0.085 | 0.52 [0.42; 0.62] | 0.05 [0.00; 0.10] | 0.43 [0.37; 0.49] | 0.50 [0.41; 0.59] | 0.05 [0.00; 0.09] | 0.45 [0.40; 0.51] | 0.78 [0.72; 0.84] |
| **aceQual** | **16** | **799**,**381** | **1**,**506**,**106** | **799**,**413** | **0.102** | **0.51 [0.45; 0.56]** | **0.05 [0.02; 0.08]** | **0.44 [0.41; 0.48]** | **0.51 [0.45; 0.56]** | **0.05 [0.02; 0.08]** | **0.44 [0.41; 0.48]** | **0.78 [0.73; 0.84]** |
| aceQuant | 17 | 799,392 | 1,506,105 | 799,426 | 0.002 | 0.38 [0.28; 0.48] | 0.11 [0.06; 0.16] | 0.50 [0.45; 0.56] | 0.58 [0.55; 0.62] | 0.00 [0.00; 0.01] | 0.41 [0.39; 0.44] | 1 (fixed) |
| ace | 15 | 799,434 | 1,506,107 | 799,464 | <0.001 | 0.45 [0.40; 0.51] | 0.05 [0.02; 0.07] | 0.50 [0.47; 0.53] | 0.45 [0.40; 0.51] | 0.05 [0.02; 0.07] | 0.50 [0.47; 0.53] | 1 (fixed) |
| aeQuantQual | 16 | 799,392 | 1,506,106 | 799,424 | 0.004 | 0.62 [0.59; 0.65] | 0 (fixed) | 0.38 [0.35; 0.41] | 0.59 [0.57; 0.62] | 0 (fixed) | 0.41 [0.38; 0.43] | 0.82 [0.77; 0.86] |
| aeQual | 15 | 799,394 | 1,506,107 | 799,424 | 0.003 | 0.60 [0.58; 0.62] | 0 (fixed) | 0.40 [0.38; 0.42] | 0.60 [0.58; 0.62] | 0 (fixed) | 0.40 [0.38; 0.42] | 0.82 [0.77; 0.86] |
| aeQuant | 15 | 799,446 | 1,506,107 | 799,476 | <0.001 | 0.56 [0.53; 0.58] | 0 (fixed) | 0.44 [0.42; 0.47] | 0.55 [0.52; 0.57] | 0 (fixed) | 0.45 [0.43; 0.47] | 1 (fixed) |
| ae | 14 | 799,446 | 1,506,108 | 799,474 | <0.001 | 0.55 [0.54; 0.57] | 0 (fixed) | 0.45 [0.43; 0.46] | 0.55 [0.54; 0.57] | 0 (fixed) | 0.45 [0.43; 0.46] | 1 (fixed) |

*Notes: EP, estimated parameters. AIC, Akaike’s Information Criterion. P, p-value for likelihood ratio test against the saturated model, h^2^, heritability, c^2^, common/shared environmental influences, e^2^, unique environmental influences, rG(sex), cross-sex genetic correlation estimated using opposite-sexed sibling pairs, 95% CI, 95% confidence intervals. Saturated refers to the saturated model. aceQuantQual is a quantitative genetic ‘ACE’ model estimating additive genetic influences (A), common environmental influences (C) and unique environmental influences (E), allowing for both qualitative and quantitative sex differences. aceQual and aceQuant are ACE models that either allow for qualitative or quantitative sex differences, respectively, whilst ace refers to an ACE model without any sex differences. The AE-models, which are variants of ACE models that exclude common environmental influences, were named similarly. Model in bold represents the most parsimonious model on the basis of AIC. Lower bounds of confidence intervals were constrained at zero, as variance components cannot be negative.*
